# The role of educational settings in the transmission chain of SARS-CoV-2 in 2020: a systematic review

**DOI:** 10.1101/2021.10.13.21264932

**Authors:** Constantine Vardavas, Katerina Nikitara, Alexander Mathioudakis, Michele Hilton-Boon, Revati Phalkey, Jo Leonardi-Bee, Anastasia Pharris, Charlotte Deogan, Jonathan E. Suk

## Abstract

**Background:** School closures have been used as a core Non pharmaceutical intervention during the COVID-19 pandemic, however the role of educational settings in COVID-19 transmission is still unclear.

**Methods:** This systematic literature review assessed studies published between December 2019 and April 1, 2021 in Medline and Embase, which included studies that assessed educational settings from approximately January 2020 to January 2021. The inclusion criteria were based on the PCC framework (P-Population, C-Concept, C-Context). The study *Population* was restricted to people 1-17 years old (excluding neonatal transmission), the *Concept* was to assess child-to-child and child-to-adult transmission, while the *Context* was to assess specifically educational setting transmission clusters.

**Results:** Fifteen studies met inclusion criteria, ranging from daycare centers to high schools and summer camps, while eight studies assessed the re-opening of schools in the 2020-2021 school year. In principle although there is sufficient evidence that children can both be infected by and transmit SARS-CoV-2 in school settings, the SAR remain relatively low -when NPI measures are implemented in parallel. Moreover, although the evidence was limited there was an indication that younger children may have a lower SAR than adolescents.

**Conclusions:** Transmission in educational settings in 2020 was minimal -when NPI measures were implemented in parallel. However, with an upsurge of cases related to variants of concern, continuous surveillance and assessment of the evidence is warranted to ensure the maximum protection of the health of students and the educational workforce, while also minimising the numerous negative impacts that school closures may have on children.

**Strengths and limitations of this study:**

- This study provides a rapid review of the peer-reviewed literature pertaining to SARS-CoV-2 transmission by children within educational settings.
- The review reflects the status quo of the previous school years (January 2020 -January 2021) due to the lag time between study implementation, peer review and publication.
- The included studies represent child-to-child transmission within the context of previous SARS-CoV-2 strains and are not directly applicable to newer variants.

## INTRODUCTION

One of the more perplexing and controversial dimensions during the first year of the COVID-19 pandemic surrounded the role of children in the transmission. Are they drivers of the pandemic, or are they merely innocent bystanders, affected in myriad ways by school closures and other physical distancing measures while not being generally at-risk of COVID-19 themselves?

Epidemiologic indicators of SARS-CoV-2 infection in children provide a complex picture regarding their potential role in the transmission chain. Systematic reviews have concluded that children and adolescents have lower susceptibility to SARS-CoV-2 infection [1, 2]. However, when infected and symptomatic, children may shed viral RNA in similar quantities to adults [3], and that younger children (under 5 years) with mild to moderate symptoms may shed even more virus than older children and adults [4]. While the proportion of asymptomatic SARS-CoV-2 infections among children in the general population is uncertain, initial data had indicated that 16% of paediatric cases in Europe in the first phase of the pandemic were classified as asymptomatic [5], while up to 90% of paediatric cases in China were deemed to be asymptomatic, mild, or moderate [6]. Moreover, it is possible that children are less often asymptomatic carriers than adults: a study of non-COVID-19-related hospitalizations in Milan identified 1% of children and 9% of adults as asymptomatic carriers of SARS-CoV-2 [7]. Meanwhile, while children are overall noted to have lower rates of severe COVID-19 cases [8], there is evidence of differing transmission dynamics between younger vs. older children [2]. There is evidence that when index cases, younger children, such as under 10 years of age, lead to lower secondary attack rates than older children and adult [9, 10].

Important potential sources of evidence surrounding the role of children in the COVID-19 pandemic come from studies situated in the community, household, healthcare or educational settings. Transmission of SARS-COV-2 has thus far been documented to be higher in household settings than in other community settings – including schools – a finding which may be potentially attributable to the individual, behavioural and contextual factors of households vs. other settings, as has been suggested elsewhere [9].

Although, at the time of writing, the more transmissible Delta variant of concern is driving SARS-CoV-2 transmission (*ref to add ECDC RRA 16-pending*) there is currently a gap in published studies looking at the transmission of Delta in school settings. However, as decisions currently need to be taken to ensure high levels of preparedness in school settings [11], the literature published thus far may have important insights to guide decision-making around school closures and re-openings, as well support decision making for mitigation measures in educational settings. This systematic literature review was conducted to assess child-to-child and child-to-adult SARS-CoV-2 transmission within educational settings and to calculate where possible the secondary attack rate (SAR) when the child is the index case.

## METHODS

### Search Strategy

This systematic literature review is reported in accordance with the Preferred Reporting Items for Systematic Reviews and Meta-Analysis (PRISMA) guidelines [12]. Relevant studies published between December 2019 and April 1, 2021 were identified by searching Medline and Embase. The following set of inclusion criteria were used to determine eligibility of the studies, which is based on the PCC framework (P-Population, C-Concept, C-Context). The study Population was restricted to people 1-17 years old (excluding neonatal transmission [13]), the *Concept* was to assess child-to-child and child-to-adult transmission when the child is the index case, while the *Context* was to assess specifically educational setting transmission clusters. Subject heading terms and free text words relating to the Population, Concept and Context terms as identified in the inclusion criteria were used to develop a comprehensive list of terms for the search strategy, from which this specific review was based. We included all studies of quantitative research, while, opinion pieces, commentaries, case reports and editorials were excluded. Mathematical modelling and simulation studies were also excluded. We additionally screened reference lists of the included articles to identify further relevant studies. The search was limited to the English language.

### Study selection

Initially, a pilot training screening process was used where 100 identical articles were screened for their eligibility independently by two reviewers to ensure consistency in screening. As a high measure of inter-rater agreement was achieved between the two reviewers during the pilot assessment (percentage agreement >90% and/ or Cohen’s Kappa >0.81), the remaining titles were randomly allocated to the two reviewers and screened for eligibility independently by them. After an initial selection of the titles, each reviewer assessed each other’s selected studies. The retrieved articles were then independently double-screened by two reviewers based on the full text of the articles.

### Data extraction

The data extraction template was piloted independently by the two reviewers on a random sample of two included studies to enable an assessment of consistency in data extraction and to identify where amendments needed to be made to the template. The remaining studies were then data extracted independently by two reviewers, and the results were double checked across the original manuscript by a third reviewer.

### Data synthesis

Characteristics of the included studies were presented in tabulated form detailing the study design, geographical location of the study, sample size, characteristics of the populations considered, setting, context, parallel implemented Non Pharmaceutical Interventions (NPI), and the findings of the study. Depending on the level of information available, infection SAR were noted. A narrative synthesis approach was applied to look systematically at the data and to describe each study categorized by the study design. Patterns in the data were identified through tabulation of results, and an inductive approach was taken to translate the data to identify areas of commonality between studies.

### Patient and Public Involvement statement

Patients or the public were not involved in the design, or conduct, or reporting, or dissemination plans of our research.

## RESULTS

### Study selection and description

A total of 5,406 studies were identified according to the specified selection criteria from Medline and Embase. After the removal of duplicates, 5,233 were screened by title/abstract, out of which 333 were assessed via full text, and 15 studies subsequently included in this review. The PRISMA flowchart showing the flow of study selection is presented in **Figure 1**.

**Figure 1.**
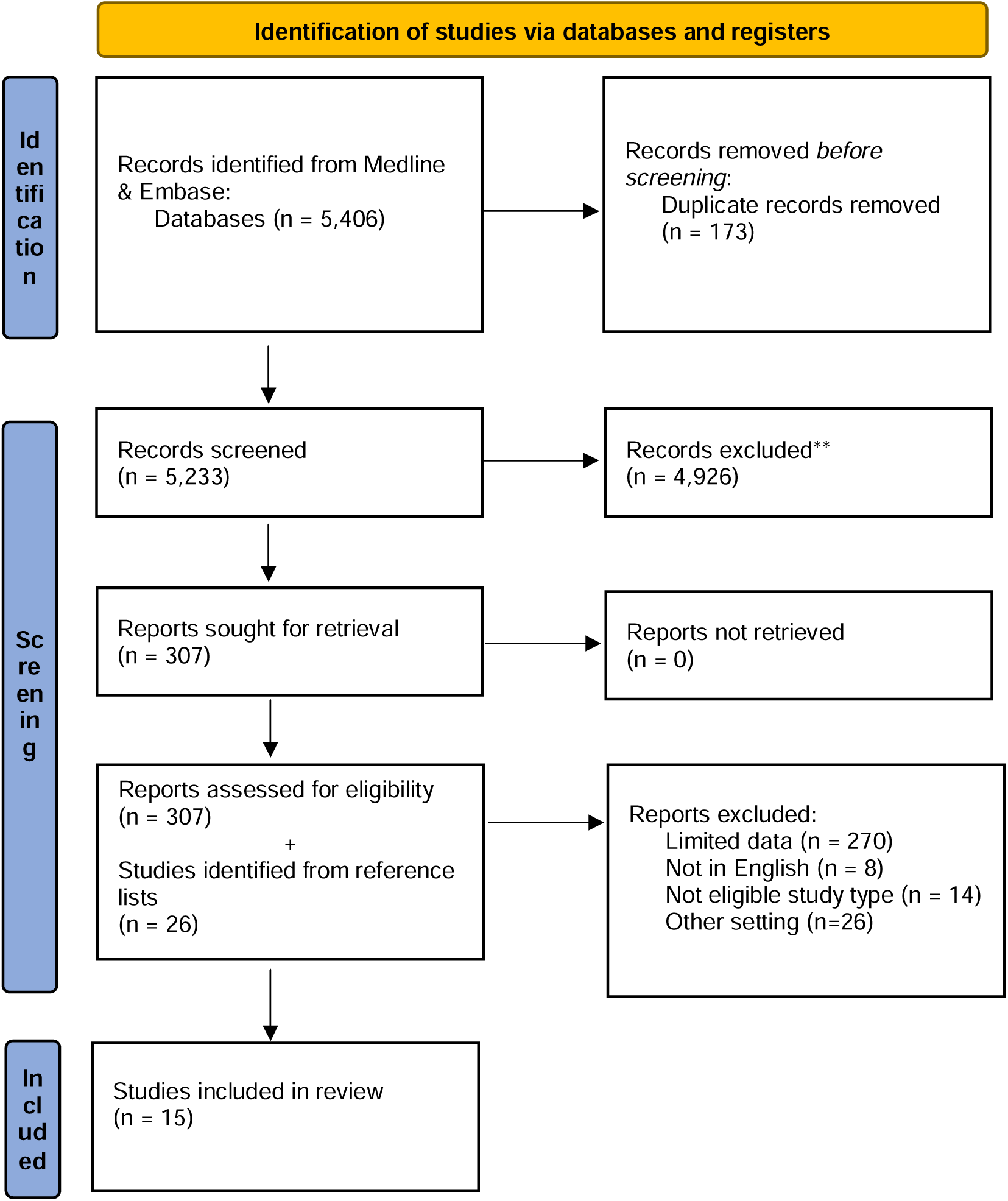
PRISMA Flowchart of study selection included in the rapid review.

Fifteen published studies were identified to address child-to-child and/or child-to-adult transmission of SARS-CoV-2. Timeframes of data collection within these studies ranged between January 2020 and January 2021. Studies from 11 countries were included (United States, South Korea, Israel, Germany, Italy, Ireland, France, Singapore, Australia, Norway, and England). A full detailed overview of the published studies is provided in **Table 1**.

**Table 1.** Studies assessing SARS-CoV-2 transmission in educational settings, reported secondary cases and parallel non pharmaceutical interventions.

| Lead Author, Year | Country | Timeframe | Age Range* | Setting | Number of symptomatic pediatric index cases | Number of asymptomatic pediatric index cases | Secondary cases in the school settings<sup>1</sup> | Parallel non pharmaceutical interventions |
| --- | --- | --- | --- | --- | --- | --- | --- | --- |
| **Child care settings** |  |  |  |  |  |  |  |  |
| Lopez et al., 2020 (18) | USA, Utah | April –July 2020 | 0.2-16 | 3 childcare facilities | 0 child (3 adults) |  | Transmission was documented from 12 secondary pediatric cases (3 asymptomatic) to at least 12/46 nonfacility contacts (confirmed or probable cases) | Quarantine for 14 days of cases + contacts; in 2 facilities: daily screening and staff members were using masks |
| Yoon et al., 2021 (24) | South Korea | February – March 2020 | 4 | 1 childcare center | 1 (information about symptoms not reported) |  | 0/190 | Adult staff wore masks, but mask wearing by children were not consistent. After the index case-patient was identified, the center was closed. All potentially exposed persons were quarantined at home for 14 days. |
| **Combined childcare-school settings** |  |  |  |  |  |  |  |  |
| Heavey et al., 2020 (14) | Ireland | March | 10-15 | Schools | 2 | 1 | 0/822 school | Exposure before school |
|  |  | 2020 |  |  |  |  | contacts<br>0/73 other<br>contacts | closure. Schools closed,<br>contacts were quarantined |
| Danis et al., 2020 **(15)** | France | January to<br>February<br>2020 | 9 | 3 schools | 1 | 0 | 0/86 school<br>contacts<br>1/6<br>hospitalised<br>contacts | Not reported |
| Yung et al., 2020 (54) | Singapore | February<br>to March<br>2020 | 2.8-15 | 3 schools | 2 | 0 | 0/42<br>symptomatic<br>contacts | Contacts were quarantined.<br>Targeted measures at the<br>school level |
| Macartney et al., 2020 **(19)** | Australia, NSW | 25 January<br>to 10 April<br>2020 | <18 | 15 schools<br>and 10<br>childcare<br>settings | 12 (information about<br>symptoms not reported) |  | 3/752 (3: 2<br>children & 1<br>adult) | Contacts were quarantined |
| Stein-Zamir et al., 2020 **(20)** | Israel | May 2020 | 12-18 | 1 high<br>school | 2 | 0 | 178/1312<br>(178: 153<br>children & 25<br>staff) | Closed spaces with poor<br>ventilation, high<br>temperatures, crowded<br>spaces and close contact<br>with no masks |
| Link-Gelles et al., 2020 (21) | USA,<br>RI | June – July<br>2020 | <18 | 666<br>educational<br>settings | 33 confirmed<br>and 19<br>probable<br>cases in 29<br>settings |  | 17 cases in<br>4/666<br>educational<br>settings with. | Class distancing, the use of<br>face masks for adults,<br>universal symptom<br>screening daily and<br>disinfection |
| Ehrhardt et al., 2020 (22) | Germany, Baden-Württemberg | May –<br>August<br>2020 | <18 | Schools<br>and<br>childcare<br>facilities | 137 (information about<br>symptoms not reported) |  | 11/>2300,<br>estimation of 1<br>secondary<br>case per<br>roughly 25<br>infectious<br>school days | Masks, social distancing,<br>hygiene, ventilation, smaller<br>class sizes, cancelled<br>activities, exclusion of sick<br>children |
| Brandal et al., 2020 (26) | Norway, Oslo and Viken counties | August –<br>November<br>2020 | 5-13 | Primary<br>schools | 13 (information about<br>symptoms not reported) |  | 3/292 (3: 2<br>children & 1<br>adult) | National guideline-based<br>infection control measures,<br>i.e. hygiene, physical<br>distancing, symptomatic<br>children to stay at home. |
|  |  |  |  |  |  |  |  | Masks not worn in schools |
| Gold et al., 2021 **(17)** | USA, Georgia | December 2020 – January 2021 | 5-13 | 8 primary schools | 1 (information about symptoms not reported) |  | 5/contacts traced not reported | Physical distancing and masks; imperfect compliance noted |
| Larosa et al., 2020 (25) | Italy, Reggio Emilia | September – October 2020 | <18 | 8 preschools, 10 primary 18 secondary | 43 | 0 | 17/1,198 (17 children & 0 adults) | Mandatory surgical masks for children except when seated and not speaking; physical distancing measures |
| Yoon et al., 2020 **(23)** | South Korea | Up to July 2020 | <18 | 6 preschools 13 primary, 6 secondary, 14 high schools | 44 (information about symptoms not reported) |  | 2/≥ 13,100 | School closure continued until 6/4/2020. Social distancing strategies and mask wearing when schools opened with rigorous contact tracing and rapid testing on any suspected cases. |
| Summer Camps |  |  |  |  |  |  |  |  |
| Pray et al., 2020 **(27)** | USA, Wisconsin | July – August 2020 | 14-24 | 1 overnight camp | 1 | 0 | 115/151 confirmed or probable cases | Documentation of a negative prearrival RT-PCR result, 7-day prearrival quarantine, and outdoor programming |
| Blaisdell et al., 2020 **(28)** | USA, Maine | June – August 2020 | 7-18 | 4 overnight camps | 0 | 1 | No secondary transmission identified | Prearrival quarantine, pre- and postarrival testing and symptom screening, cohorting, use of face coverings, physical distancing, enhanced hygiene measures, cleaning and disinfecting, and maximal outdoor programming |

1: Measured from the date of last contact; 2: Probable cases; \*Except when the age refers to only 1 pediatric case and age range is n/a

### Studies assessing outbreaks in Educational Settings

Heavey et al. [14] conducted a case study in order to explore the role of transmission among children in the school setting in the Republic of Ireland, before school closure. Three pediatric index cases of COVID-19 with a history of school attendance were detected with 895 contacts. Child-to-adult transmission or child-to-child transmission was not reported in this study. Similarly Danis et al. [15] presented the contact tracing results of a nine-year-old child in France, who visited 3 different schools the first days of symptom appearance. There was no evidence of secondary transmission in any of the school contacts. Moreover, Yung et al. traced three COVID-19 cases (2 pediatric and 1 adult) in three different educational settings, and the results were negative, as were the tracing of close contacts of a preschool case in S Korea [16]. Gold et al, in early 2020 had also indicated the possibility of educators playing a role in school transmission as identified through the assessment of a transmission cluster in primary (elementary) schools in the US [17] while Lopez et al assessed three COVID-19 outbreaks in child care facilities in Utah, during April 1–July 10, 2020 and noted that SARS-CoV-2 Infections among young children acquired in child care settings were transmitted to their household members [18].

One study from New South Wales, Australia presented an overview of COVID-19 cases and transmission in schools. In a total number of 15 schools and 10 Early Childhood Educational and Care Settings, 27 index cases were identified, among which 12 were children and 15 staff members. Secondary transmission was noted only in four out of 25 educational settings, where 2 children and 1 adult secondary cases were detected after the tracing of 752 contacts [19].

### Studies assessing the re-opening of schools and summer camps

Eight studies reported on the regional evidence after the re-opening of schools. A school outbreak in Israel after reopening of schools in May 2020 was described by Stein-Zamir et al. The outbreak assessment was initiated by two pediatric COVID-19 cases that were not epidemiologically related. The results showed that 153/1161 students and 25/151 staff members tested positive for COVID-19. However, this outbreak was attributed to crowded classes, combined with the exemption of facemasks and the use of air-condition due to an extreme heatwave [20]. On the contrary, a study by Link-Gelles et al., in Rhode Island, USA. among 666 child care programs that reopened on 1 June, 2020 after a 3-month closure revealed 52 confirmed and probable cases (33 confirmed cases), of which 30 were among children and 22 among adults. Secondary transmission for 10 cases was noted in only 4/666 childcare programs, which was attributed to class distancing, the use of face masks for adults, universal symptom screening daily and disinfection [21]. The regional reopening of schools in Germany in May 2020 was assessed by Ehrhardt et al., who noted that child-to-child transmission in schools/childcare facilities appeared very uncommon, with an estimated six of the identified 137 cases that had attended school to have led to a secondary transmission overall to 11 additional pupils [22]. While two additional studies from S Korea by Yoon et al., indicated that upon the return of children to school in May-June 2020, no indication of secondary transmission was noted in kindergarten children, middle school or high schools, while in primary school only two cases of secondary transmission was noted [23, 24]. The reopening of schools in September 2020 in Italy was not associated with elevated SAR, which reached 3.8% overall, 0% in preschool, 0.38% in primary and 6.46% in secondary schools, however these percentages included both adult and child cases [25]. Brandal et al., assessed the transmission of COVID-19 in school settings in Norway between August-November 2020 and identified minimal child-to-child (0.9%, 2/234) and child-to-adult (1.7%, 1/58) transmission [26].

Summer educational camps are presented separately, as close proximity between students is not only noted within school hours but throughout the day and night due to additional extra curriculum activities and close sleeping proximity. Two studies assessed secondary transmission within summer educational camps, with striking differences. Pray et al identified a rapid transmission of SARS-CoV-2 at an overnight retreat where adolescents and young adults aged 14–24 years had prolonged contact and shared sleeping quarters, where one index case/child led to the infection of 76% of attendees [27]. On the contrary Blaisdell in four overnight camps noted no indication of secondary transmission following the isolation of the paediatric index case and quarantine of their cohort, indicating the importance of the implementation of NPI to reduce COVID-19 transmission [28].

### Secondary attack rates of COVID-19 transmission in educational settings

**Table 2** presents the SAR extracted from the studies, ranging from 0 to 76%, depending on the setting, the timeframe and the implementation of NPI. With the exception of the study by Pray et al., [27] within the context of summer camps in which a high transmission rate (76%) was noted, in all studies within the context of school settings, the reported SARs were minimal. Age differentiations were noted, for instance in the study by Larosa et al., across 36 schools in northern Italy, who identified an overall SAR of 3.2%, reaching 6.6% in middle and high schools and 0.38% in primary schools.

**Table 2.** Studies that assessed the secondary attack rate (SAR), when children are the index case within educational settings.

| Lead Author, Year | Country | Timeframe | SAR |
| --- | --- | --- | --- |
| Heavey et al., 2020 **(14)** | Ireland | March 2020 | 0 |
| Danis et al., 2020 **(15)** | France | January to February 2020 | School: 0/86, Community: 0/80, Hospitalised: 1/6 |
| Yung et al., 2020 **(54)** | Singapore | February to March 2020 | 0/42 |
| Macartney et al., 2020 **(19)** | Australia, NSW | 25 January to 9 April, 2020 | All settings, all child case to child contacts 0.3% (2/649)<br>All settings, all child case to staff member contacts 1.0% (1/103),<br>Child close contacts 28.0% (7/25) |
| Stein-Zamir et al., 2020 **(20)** | Israel | May 2020 | 178 / 1312 |
| Heavey et al., 2020 **(14)** | USA, Rhode Island | 1 June- 31 July, 2020 | n/a |
| Pray et al., 2020 **(27)** | United States, Wisconsin | July-August 2020 | 115/151 (76%) |
| Blaisdell et al., 2020 **(28)** | United States, Maine | June-August 2020 | 0 |
| Lopez et al., 2020 **(18)** | USA, Utah | April-July 2020 | n/a |
| Ehrhardt et al., 2020 (22) | Germany, Baden-Württemberg | 25 May - 5 August 2020 | estimation of one secondary case per roughly 25 infectious school days |
| Brandal et al., 2020 (26) | Norway, Oslo and Viken counties | 28 August - 11 November 2020 | child 2/234 (0.9%),<br>adult 1/58 (1.7%) |
| Gold et al., 2021 (17) | United States, Georgia | 1 December 2020 - 22 January 2021 | n/a |
| Larosa et al., 2020 (25) | Italy | 1 September -15 October 2020 | 38/994 (3.82%) overall<br>0.38% in primary schools (1/266)<br>6.46% in secondary schools (37/572) |
| Yoon et al., 2021 (24) | South Korea | 27/2-16/3/2020 | 0 |
| Yoon et al., 2020 (23) | Korea | up to 31/7/2020 | 2/≥ 13,100 |

## DISCUSSION

This study provides a rapid review of the peer-reviewed literature pertaining to SARS-CoV-2 transmission by children within educational settings, a topic which is a crucial input to assessments of the role of school settings in COVID-19 transmission. The literature appraised in this review provides sufficient evidence that children can both be infected by and transmit SARS-CoV-2 in school settings, however the SAR remained relatively low within the studies assessed by our review, reflecting primarily schools in 2020. Our results with regards to educational settings are in line with population based studies published after the cut-off of this review, in which SARS-CoV-2 outbreaks were uncommon in educational settings [29] in England [30], Canada [31] and in Utah, [32], Missouri [33] and New Jersey, USA [34], during similar periods.

During the first waves of the COVID-19 pandemic, the vast uncertainty surrounding the epidemiology of SARS-CoV-2 led many countries globally to include school closure concomitant with other NPIs for reducing COVID-19 transmission. Within our review there were limited cases in the assessed studies in which a child index case was responsible for extensive secondary transmission in schools, with the notable exception of an outbreak in Israel (which was associated with dense spacing, lack of the use of facemasks and closed spaces with poor ventilation) and secondary transmission within summer educational camps, where prolonged exposure between case-contact pairs is expected [27]. This finding is supported by data from a large population based study assessing transmission dynamics that identified that patterns of enhanced transmission risk in similar age pairs were strongest among children aged 0 to 14 years [2].

On the contrary, evidence from studies that note a very small number of cases after school reopenings the authors attribute to the strict NPIs implemented including the use of face masks, physical distancing, screening for symptoms and classroom disinfection. Close proximity between students was linked to elevated transmission rates in both school settings and educational camps [20, 27], while adult educators have also been noted to play a role in school transmission [17].

Modelling studies using various assumptions of infectivity from the first 3-4 months of the pandemic [35-41], have previously assessed the role of school closure, and overall indicated that school closure is associated with a reduction in the number of cases, hospitalisations and ICU admissions, with the effect of school closure dependent on the transmission rate, and the duration of school closure. Within this context age is noted to be a crucial aspect, as recent modelling studies from the Netherlands indicated that contact restrictions within the age group of 10-20 years old caused a slightly more significant reduction in Re compared to 5-10 years old [54]. The same study also assessed the impact of reducing school contacts in pandemic progression and showed that if complete school closure were implemented after the summer holidays, R would be reduced by 10%, however, if school closure was enacted in November, after implementing a partial lockdown since August, it could further decrease R by 16%. Another recent European study that assessed school closure, based on the population of two large cities of Norway, Oslo and Tromso, indicating that a controlled and gradual school re-opening would only have a slight increase in the reproduction number of less than 0.25, and probably in the range between 0.10 and 0.14, which would not substantially affect the infection rates [55].

While school closure may reduce SARS-CoV-2 transmission as noted above, the societal and economic impacts of prolonged school closure are noteworthy, as they may impact the availability of the healthcare workforce [37, 42] and may also have negative effects on children through the interruption of the educational learning, social isolation, increased exposure to domestic violence, and rise in dropout rates [43]. Furthermore, the impact of school closures has been noted to impact significantly also special education [44], while research performed within the context of the COVID-19 pandemic has identified that contextual factors of particular relevance during school closures had negative impacts on student wellbeing [45]. In light of the above, policy makers need to be aware of the cost/benefit in each setting when considering school closures as a NPI [46].

Transmission of SARS-COV-2 has been noted to be higher in household settings than other community settings, including schools, a finding which may be potentially attributable to the individual, behavioural and contextual factors of the household vs. other settings, which may support transmission dynamics [47]. Direct evidence showing children as a source of transmission is scarce and largely based on small studies or studies investigating few paediatric cases, however the results presented here concur with other and previous systematic reviews that have summarised the evidence on the role of children in SARS-CoV-2 transmission [48-50].

There are important limitations to this study that may impact the direct implications for decision-making. As we assessed peer-reviewed evidence published in two biomedical databases, it inherently reflects the status quo of the interim of the previous school years (January 2020 -January 2021) due to the lag time between study implementation, peer review and publication. A further limitation of this report refers to the fact that these studies represent child-to-child transmission within the context of previous SARS-CoV-2 strains and are not directly applicable to newer and more transmissible variants, such as the SARS-CoV-2 Delta (B.1.617.2) variant of concern. Finally, the included studies reflect a broad geographical and temporal range and are limited in comparability due to varying factors such as: background levels of community SARS-CoV-2 transmission; enrolment strategies and varying NPI policies which in turn depends highly on the geographical region and the socioeconomic context, while accountability to government and political stability were found to exert influence [51]. Hence in light of the above, supporting educators and parents in the implementation of NPIs is important as population based studies have indicated that adults concerned about the impact of COVID-19 on their children’s education were more likely to practice personal protective measures and social distancing [52].

## CONCLUSIONS

The findings presented here provide an assessment of the published peer-reviewed evidence on transmission in educational settings during 2020, in which transmission was minimal -when NPI measures were implemented in parallel. However, with an upsurge of cases related to new variants of concern, notably Delta, continuous surveillance and assessment of the evidence is warranted to ensure the maximum protection of the health of students and the educational workforce, while also minimising the numerous negative impacts that school closures may have on children. Where schools remain open, in-school NPI measures should be continually refined according to new knowledge according to the epidemiologic context, taking into account levels of community SARS-CoV-2 transmission, information on the severity of the Delta variant, and vaccination coverage levels among eligible students, which includes children over 12 in many jurisdictions, at the time of writing [53].

## Data Availability

No data have been produced. It is a systematic review.

## Acknowledgments

We would like to thank Katerina Papathanasaki, Chrysa Chatzopoulou, Konstantinos Skouloudakis and Ioanna Lagou for their assistance in data archiving and report preparation. We acknowledge the bravery and dedication of educational professionals everywhere during these most challenging times.

## Availability of data and material

Not applicable

